# Development of an automated, imaging-based preoperative screening model for early identification of malnutrition in an abdominal surgery cohort

**DOI:** 10.64898/2026.06.08.26355187

**Authors:** Victoria Gershuni, Raheema A. Damani, Shubha Vasisht, Rakesh Sharma, James Rowe, Charlene Compher, Jeffrey Duda, Hersh Sagreiya, Rachel Kelz, Hongzhe Li, Gregory Tasian, Scott Damrauer, Gary D. Wu, Walter R. Witschey

**Author notes:** Correspondence: Victoria Gershuni, MD MTR MSGM, 3400 Spruce Street, 4 Silverstein, Philadelphia, PA 19104.

## Abstract

**Background:** Clinical malnutrition affects one in five abdominal surgery patients and increases postoperative complications and mortality. Current screening occurs after admission, closing the window for preoperative nutritional intervention. No objective, scalable preoperative screening tool exists.

**Objective:** To determine whether automated volumetric CT-based body composition analysis improves preoperative identification of surgical patients at risk for clinical malnutrition compared to clinical variables or single-slice imaging alone.

**Methods:** Retrospective cohort study of adults undergoing elective abdominal surgery at a quaternary academic medical center (2018–2021) with a preoperative CT scan within 90 days and complete nutrition assessment. Clinical malnutrition was diagnosed by a registered dietitian using ASPEN/AND criteria. Three sex-stratified Elastic Net models were compared: (1) base clinical variables; (2) base plus L3 single-slice skeletal muscle index and attenuation; and (3) base plus comprehensive 3D volumetric quantification of five muscle groups and two fat depots. Discrimination (AUROC), calibration (Brier score), and clinical utility (decision curve analysis) were assessed via 10-fold cross-validation.

**Results:** Among 1,143 patients (52.4% female; mean age 60.5 years), 231 (20.2%) were diagnosed with malnutrition. Malnourished patients had significantly higher complication rates (36.4% vs. 15.4%, *p*<0.001) and prolonged length of stay (45.9% vs. 16.4%, *p*<0.001). Critically, 27.2% of malnourished patients were not flagged as at-risk by the standard Malnutrition Screening Tool. The volumetric model (Model 3) achieved the highest discrimination (males: AUROC 0.808; females: 0.794) and best calibration (males: Brier 0.129; females: 0.124), significantly outperforming both the base model (males: *p*=0.004; females: *p*<0.001) and L3 model (males: *p*=0.019; females: *p*<0.001). L3 features modestly improved discrimination but paradoxically worsened calibration — an effect corrected by volumetric features. Sex-specific risk profiles differed markedly, with ASA classification dominating female models and demographic factors dominating male models.

**Conclusions:** Automated volumetric CT body composition analysis significantly improves preoperative malnutrition risk identification, with sex-stratified models revealing distinct risk profiles. Leveraging imaging already obtained for surgical planning, this approach opens a preoperative window for nutritional intervention that current practice fails to utilize.

## Introduction

Malnutrition is highly prevalent among abdominal surgery patients yet remains largely unrecognized in the preoperative setting, representing a missed opportunity for intervention. The current malnutrition screening paradigm for most elective surgical patients occurs after hospital admission ^1^ when patients have already undergone surgery and the window for preemptive nutritional optimization has closed. This timing issue is critical: malnourished abdominal surgery patients experience a threefold increase in complications and fivefold increase in mortality compared to well-nourished patients.^2–4^ Despite malnutrition being a modifiable risk factor, the lack of objective, automated screening tools deployable in outpatient surgical clinics perpetuates late diagnosis and suboptimal outcomes.

Current preoperative nutritional assessment relies heavily on subjective measures including body mass index (BMI) and patient-reported weight loss, neither of which adequately captures physiologic reserve or surgical readiness.^5,6^ BMI fails to distinguish between muscle and fat mass, while patient-perceived weight loss is particularly unreliable in patients with obesity or those experiencing weight-stable changes in body composition (loss of muscle with concurrent fat gain).^6^ Muscle mass assessment, recognized as a key component of malnutrition diagnosis, represents a patient’s physiologic reserve to respond to surgical stress yet is not routinely measured in clinical practice.^7^ The Academy of Nutrition and Dietetics and the American Society for Parenteral and Enteral Nutrition emphasize the underappreciated role of low muscle mass in malnutrition management, but practical tools for measuring muscle preoperatively remain limited.^7–9^

Computed tomography (CT) provides objective quantification of body composition and has been extensively studied in research settings. Single-slice axial imaging at the third lumbar vertebrae (L3) can measure cross-sectional skeletal muscle area (skeletal muscle index (cm^2^/m^2^), SMI) and average attenuation (skeletal muscle radiation attenuation (HU), SMRA) to identify sarcopenia and myosteatosis, respectively. Retrospective studies demonstrate associations between these features and adverse surgical outcomes,^10–17^ but translation to clinical practice has been hindered by the need for manual segmentation requiring highly trained personnel, lack of integration into clinical workflows, and high costs. Several groups have developed automated approaches for L3 muscle segmentation using convolutional neural networks and deep learning.^18–22^ Our group recently published a volumetric algorithm that extends beyond single-slice measurements to quantify individual muscle groups and fat depots across the entire abdomen.^23,24^ While these technological advances enable scalable measurement, the first question remains: can automated body composition analysis effectively screen for malnutrition risk before surgery?

We recently demonstrated significant sex-specific associations between CT-derived body composition features and clinical malnutrition diagnosis in abdominal surgery patients.^25^ Females exhibited distinct patterns of muscle quality deterioration (myosteatosis) associated with malnutrition, while males showed different patterns of muscle size reduction and visceral fat depletion. These findings highlight that screening models must account for sex-specific differences rather than applying uniform cutoffs. Building on these associations, the current study advances to the predictive question: can preoperative CT-derived body composition features predict which patients will be diagnosed with malnutrition, effectively serving as an automated screening mechanism?

In this study, we develop and compare three predictive modeling approaches to identify preoperative malnutrition risk. First, we establish a base model using standard clinical variables routinely available in the preoperative setting (age, race, weight, comorbidities). Second, we augment this base model with L3 SMI and SMRA– the traditional single-slice body composition measures most reported in the literature—to determine whether adding conventional imaging features improves prediction beyond clinical variables alone. Third, in conjuction with clinical and L3 features, we incorporate our comprehensive volumetric body composition assessment, including measurements of individual muscle groups and fat depots, to evaluate whether detailed three-dimensional analysis provides incremental predictive value over both clinical variables and single-slice measurements. By comparing model performance across these three approaches in sex-stratified analyses, we can determine whether automated, comprehensive body composition analysis justifies its integration into preoperative screening workflows. Preoperative nutrition support has been demonstrated to improve outcomes post-operatively. ^26–29^ Therefore, we secondly hypothesize that predictive models incorporating comprehensive, automated body composition analysis from routine preoperative CT scans will significantly improve identification of patients at risk for malnutrition compared to clinical variables alone or traditional single-slice measurements, enabling timely nutritional intervention and optimization.^30^

## Methods

### Study Population

This retrospective study included adult patients at a single quaternary care institution (2018-2021) who underwent elective abdominal surgery with routine preoperative CT scan within 90 days, as previously described.^25,31^ Briefly, data were prospectively collected for the American College of Surgeons National Surgery Quality Improvement Program (ACS NSQIP) registry for five abdominal operations (pancreatectomy, hepatectomy, colectomy, proctectomy, and ventral hernia repair).^32^ Clinical, demographic, and comorbidity data were abstracted from the electronic health record, PennChart (Epic Systems Corporation, Verona, WI).

### Nutrition Outcomes

The primary outcome was clinical malnutrition diagnosed by a Registered Dietitian using the Academy of Nutrition and Dietetics - ASPEN Indicators of Malnutrition (AAIM) criteria following a standardized two-step nutrition care process, as previously described.^33^ Patients with missing screening assessment and no diagnosis of clinical malnutrition were treated as having no risk for malnutrition in the primary analysis (males: *n* = 50 missing; females: *n* = 64 missing).

### Adverse Surgical Outcomes

Secondary outcomes included any post-operative complication (composite of infectious, respiratory, cardiac, renal, and/or thromboembolic events from NSQIP data) and prolonged length of stay (>75^th^ percentile for the procedure-specific length of stay). Emergent operations were excluded.

### CT-Derived Body Composition Assessment

CT imaging protocols and automated body composition quantification using deep learning have been previously described. ^24,34,35^ Briefly, two-dimensional (2D) L3 single-slice measures (skeletal muscle index [SMI] and skeletal muscle radiation attenuation [SMRA]) were obtained using licensed software (DAFS v3.6, Voronoi Health Analytics). Three-dimensional (3D) volumetric quantification of five muscle groups (psoas, erector spinae, quadratus lumborum, lateral abdominals, and rectus abdominus) and two fat depots (visceral and subcutaneous) measured both volume index (cm^3^/m^2^) and attenuation (HU) across T12-L5 vertebral levels (Figure 1). All volumetric measurements were standardized using sex-specific z-score normalization. The study design is shown in Figure 1. A complete list of imaging features is provided in Supplementary Table 1.

**Figure 1:**
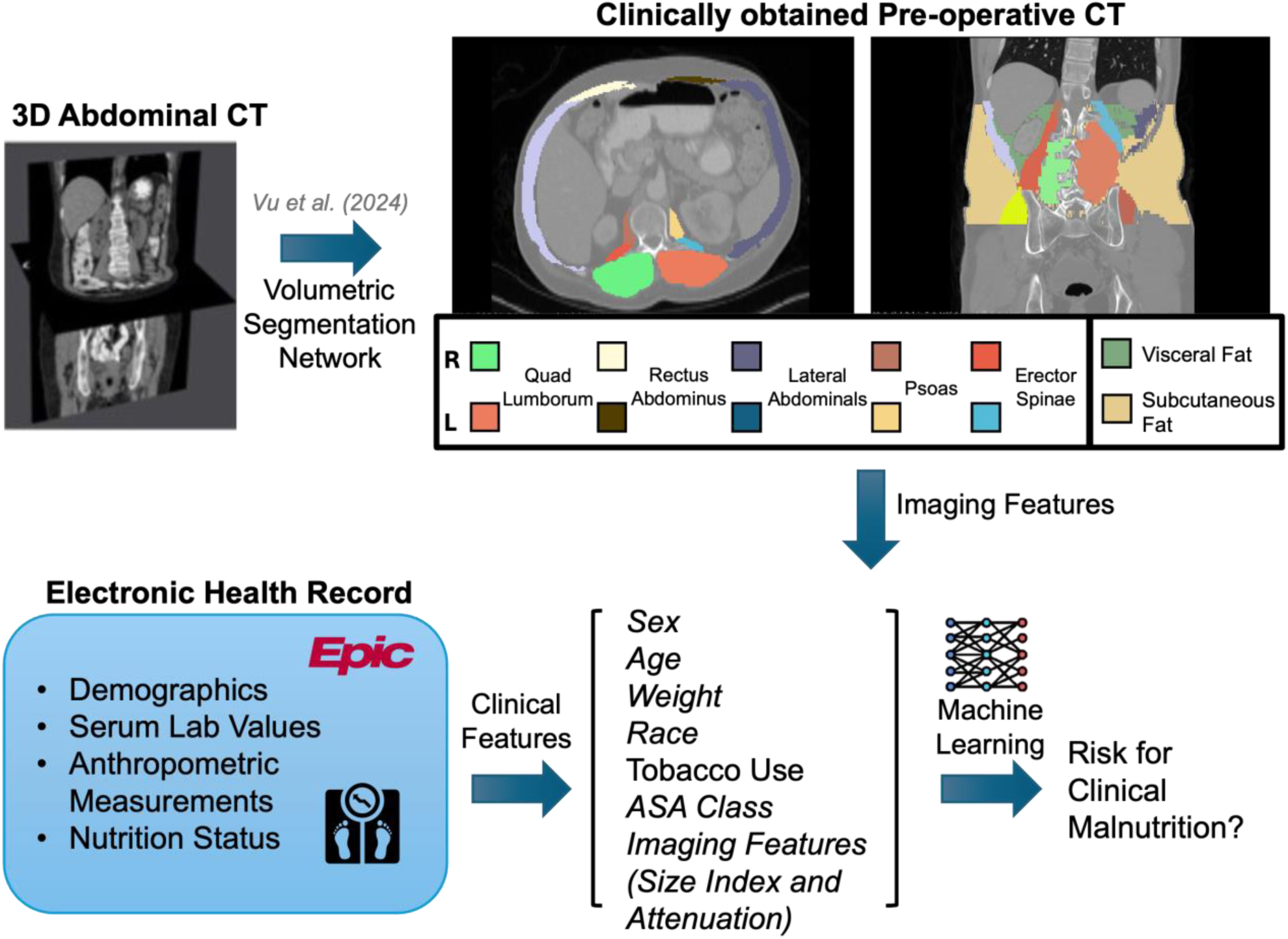
An automated machine learning algorithm for volumetric segmentation of size and attenuation characteristics for individual muscle groups and fat depots was applied to clinically obtained CT scans to extract imaging derived body composition features. Imaging features include size (height-adjusted volume, cm^3^/m^2^) and attenuation (Hounsfield Unit, HU) for five muscle groups (psoas, quadratus lumborum, rectus abdominus, lateral abdominals, erector spinae, and two fat depots- visceral and subcutaneous. Predictive models were developed to screen for malnutrition in the preoperative time period were developed incrementally, utilizing clinical data from the electronic health record and imaging derived features.

### Model Development and Validation

#### Predictor Variables

We developed sex-stratified predictive models using three feature sets to enable systematic comparison of predictive value:

1. Base Model: Clinical and demographic variables with p<0.05 on univariate analysis (age, race, weight, tobacco use, ASA classification).
2. Base + L3 Model: Base clinical variables plus traditional L3 axial single-slice imaging features (L3 SMI and SMRA).^36–38^
3. Base + L3 + Volumetric Model: Base clinical variables plus both L3 single-slice measurements and comprehensive 3D volumetric body composition features (14 imaging variables including volume index and attenuation for all five muscle groups and two fat depots). This model incorporates an incremental approach to include all variables together in a single model to evaluate their complementary impact.

#### Modeling Approach

Elastic Net logistic regression was employed for all models, combining L1 (Lasso) and L2 (Ridge) penalties to handle correlated features and perform automated feature selection, resulting in parsimonious models. ^39^ Separate models were developed for males and females.

#### Model Performance

Model discrimination and calibration were assessed using complementary metrics. Area under the receiver operating characteristic curve (AUROC) with 95% confidence intervals quantified the model’s ability to discriminate between patients with and without malnutrition, with values calculated using five repeats of ten-fold cross-validation. The DeLong test compared AUROC performance between models. Model calibration—the agreement between predicted probabilities and observed outcomes—was assessed using multiple approaches. The Brier score was calculated as the mean squared difference between predicted probabilities and actual outcomes across all predictions.^40^ Brier scores range from 0 to 1, with lower values indicating better calibration; a score of 0 represents perfect prediction, while 0.25 represents the expected score of a non-informative model for a binary outcome with 25% prevalence (matching our population). Brier scores were calculated using the same cross-validation framework as AUROC to ensure comparable assessment. While AUROC measures the model’s ability to rank patients by risk (discrimination), Brier score assesses the accuracy of the predicted probabilities themselves (calibration), which is critical for clinical decision-making when precise risk estimates are needed to guide intervention intensity. Feature importance was assessed to identify key contributors and sex-specific patterns.^41^

#### Clinical Application Tools

Decision curve analysis (DCA) assessed clinical net benefit probability thresholds to determine the utility of screening versus standard care.

### Statistical Analysis

Descriptive statistics summarize the cohort, comparing patients with and without clinical malnutrition. Continuous variables were analyzed using Student’s t-test or Mann-Whitney U test; categorical variables using Chi-square or Fisher’s exact test. All analyses were performed using *tidymodels* package in R version 4.1.2 (R Project for Statistical Computing, Vienna, Austria) or Python 3.8. Statistical significance was set at p<0.05. This study complied with TRIPOD-AI guidelines and was deemed exempt by the institutional review board.

## Results

### Study Cohort and Malnutrition Prevalence

The cohort (n=1,143; 47.6% male, 75% white) had a mean age of 60.5 ± 14.6 years old and BMI of 28.7 ± 7.1 kg/m2 (Table 1). The most common procedure was colectomy (44%), followed by ventral hernia repair (20%), pancreatectomy (18%), hepatectomy (14%), and proctectomy (4%). Clinical malnutrition was diagnosed in 231 patients (20.2%), with varying prevalence between pancreatectomy (78 of 204, 38.2%), colectomy (120 of 504, 23.8%), proctectomy (11 of 51, 21.6%), hepatectomy (14 of 155, 9.0%), and ventral hernia repair (8 of 229, 3.5%, p<0.001, Figure 2 and Table 2). Patients with clinical malnutrition had significantly higher rates of post-operative complications (36.4% vs. 15.4%, p<0.001) and prolonged length of stay (45.9% vs. 16.4%, p<0.001) compared to well-nourished patients (Table 1). Complete demographic and clinical characteristics stratified by sex have been previously reported. ^25^

**Figure 2:**
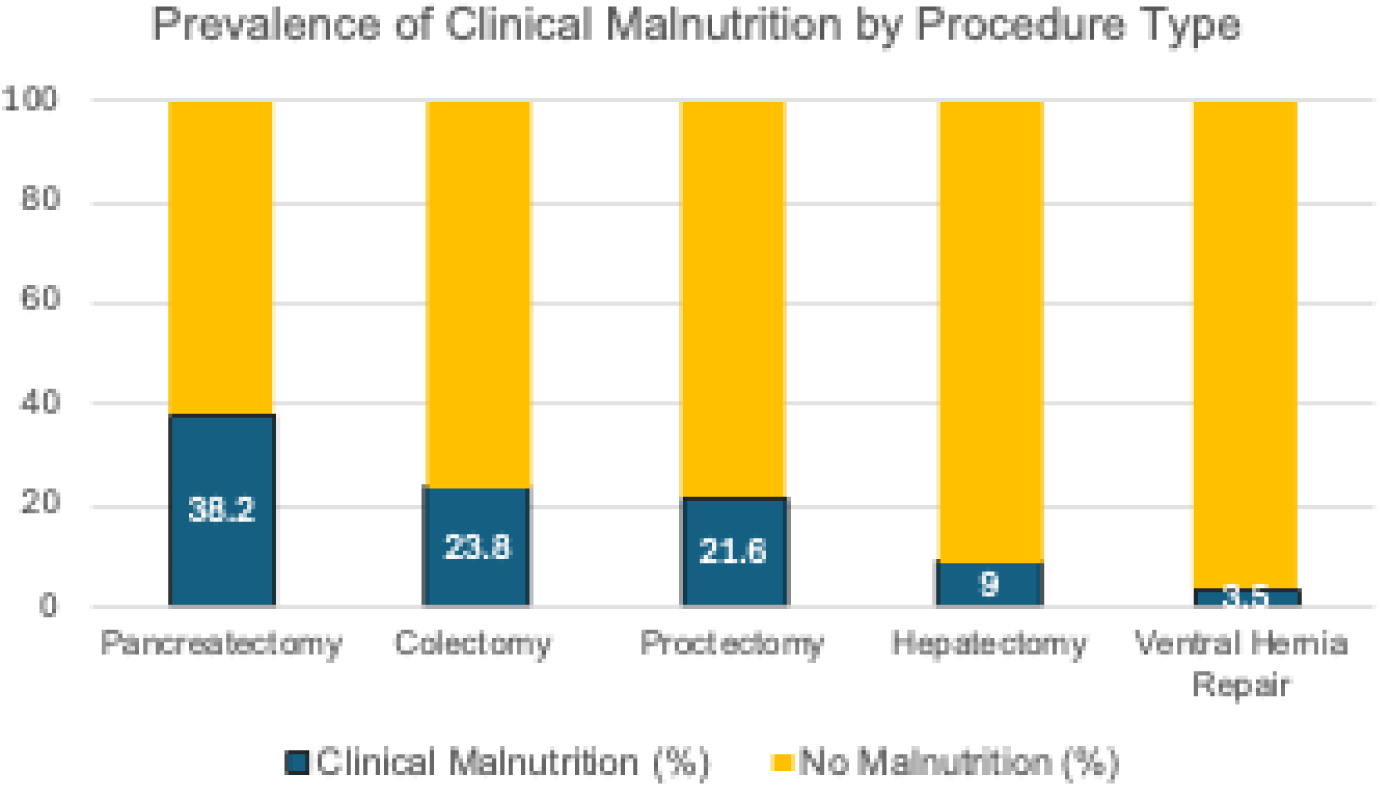
Prevalence of patients with diagnosis of clinical malnutrition in different abdominal surgery procedure types.

**Table 1.**
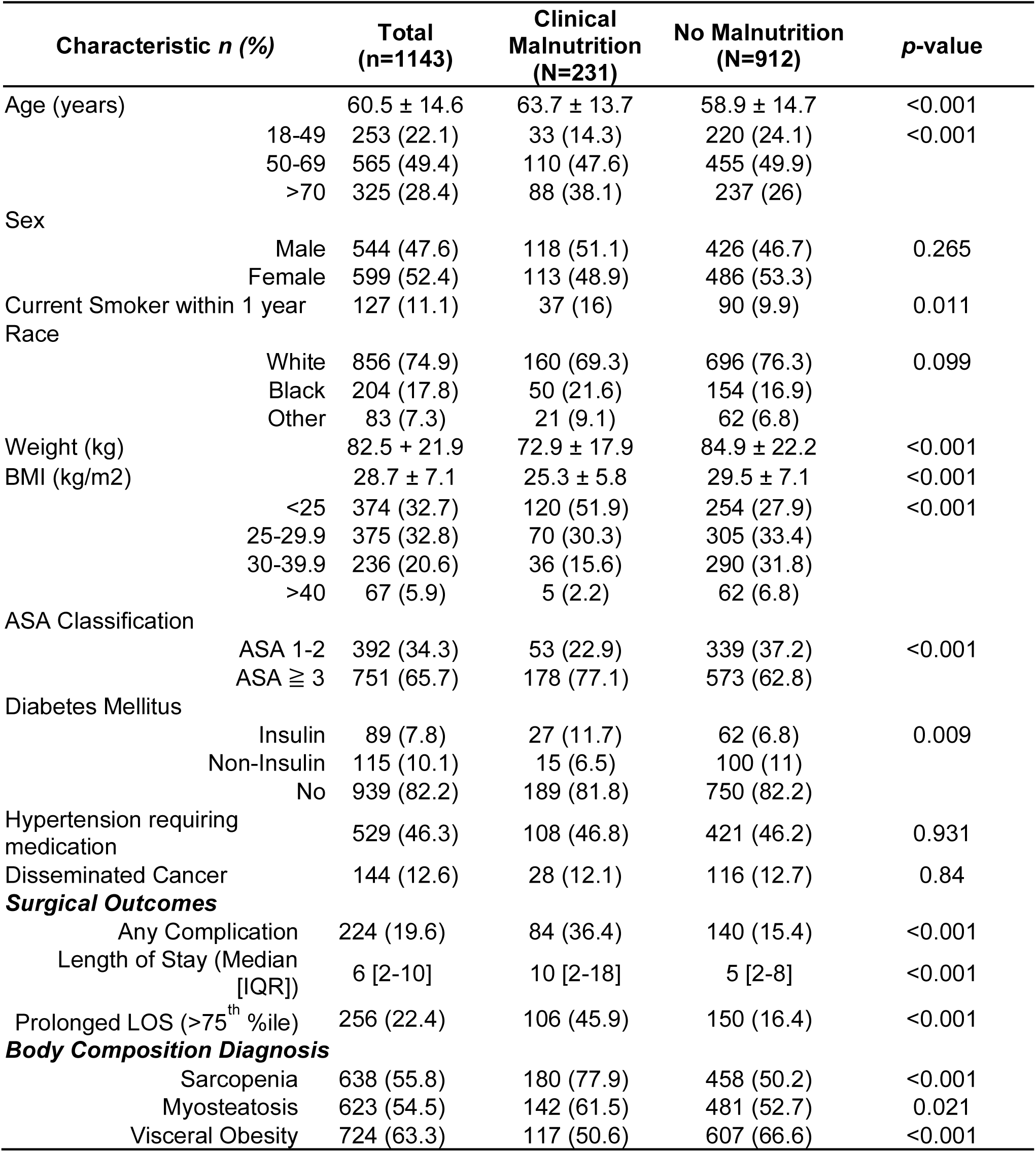
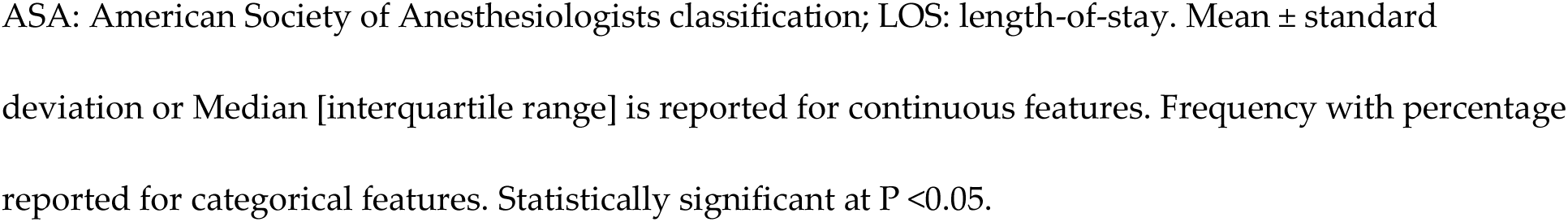
Descriptive Statistics Comparing Patients with Diagnosis of Clinical Malnutrition to Patients without Malnutrition in Study Population.

| Characteristic *n* (%) | Total<br>(*n*=1143) | Clinical<br>Malnutrition<br>(*N*=231) | No Malnutrition<br>(*N*=912) | *p*-value |
| --- | --- | --- | --- | --- |
| Age (years) | 60.5 ± 14.6 | 63.7 ± 13.7 | 58.9 ± 14.7 | <0.001 |
| 18-49 | 253 (22.1) | 33 (14.3) | 220 (24.1) | <0.001 |
| 50-69 | 565 (49.4) | 110 (47.6) | 455 (49.9) |  |
| >70 | 325 (28.4) | 88 (38.1) | 237 (26) |  |
| Sex |  |  |  |  |
| Male | 544 (47.6) | 118 (51.1) | 426 (46.7) | 0.265 |
| Female | 599 (52.4) | 113 (48.9) | 486 (53.3) |  |
| Current Smoker within 1 year | 127 (11.1) | 37 (16) | 90 (9.9) | 0.011 |
| Race |  |  |  |  |
| White | 856 (74.9) | 160 (69.3) | 696 (76.3) | 0.099 |
| Black | 204 (17.8) | 50 (21.6) | 154 (16.9) |  |
| Other | 83 (7.3) | 21 (9.1) | 62 (6.8) |  |
| Weight (kg) | 82.5 ± 21.9 | 72.9 ± 17.9 | 84.9 ± 22.2 | <0.001 |
| BMI (kg/m<sup>2</sup>) | 28.7 ± 7.1 | 25.3 ± 5.8 | 29.5 ± 7.1 | <0.001 |
| <25 | 374 (32.7) | 120 (51.9) | 254 (27.9) | <0.001 |
| 25-29.9 | 375 (32.8) | 70 (30.3) | 305 (33.4) |  |
| 30-39.9 | 236 (20.6) | 36 (15.6) | 290 (31.8) |  |
| >40 | 67 (5.9) | 5 (2.2) | 62 (6.8) |  |
| ASA Classification |  |  |  |  |
| ASA 1-2 | 392 (34.3) | 53 (22.9) | 339 (37.2) | <0.001 |
| ASA ≥ 3 | 751 (65.7) | 178 (77.1) | 573 (62.8) |  |
| Diabetes Mellitus |  |  |  |  |
| Insulin | 89 (7.8) | 27 (11.7) | 62 (6.8) | 0.009 |
| Non-Insulin | 115 (10.1) | 15 (6.5) | 100 (11) |  |
| No | 939 (82.2) | 189 (81.8) | 750 (82.2) |  |
| Hypertension requiring medication | 529 (46.3) | 108 (46.8) | 421 (46.2) | 0.931 |
| Disseminated Cancer | 144 (12.6) | 28 (12.1) | 116 (12.7) | 0.84 |
| ***Surgical Outcomes*** |  |  |  |  |
| Any Complication | 224 (19.6) | 84 (36.4) | 140 (15.4) | <0.001 |
| Length of Stay (Median [IQR]) | 6 [2-10] | 10 [2-18] | 5 [2-8] | <0.001 |
| Prolonged LOS (>75<sup>th</sup> %ile) | 256 (22.4) | 106 (45.9) | 150 (16.4) | <0.001 |
| ***Body Composition Diagnosis*** |  |  |  |  |
| Sarcopenia | 638 (55.8) | 180 (77.9) | 458 (50.2) | <0.001 |
| Myosteatosis | 623 (54.5) | 142 (61.5) | 481 (52.7) | 0.021 |
| Visceral Obesity | 724 (63.3) | 117 (50.6) | 607 (66.6) | <0.001 |

ASA: American Society of Anesthesiologists classification; LOS: length-of-stay. Mean ± standard deviation or Median [interquartile range] is reported for continuous features. Frequency with percentage reported for categorical features. Statistically significant at $P < 0.05$ .

**Table 2.**
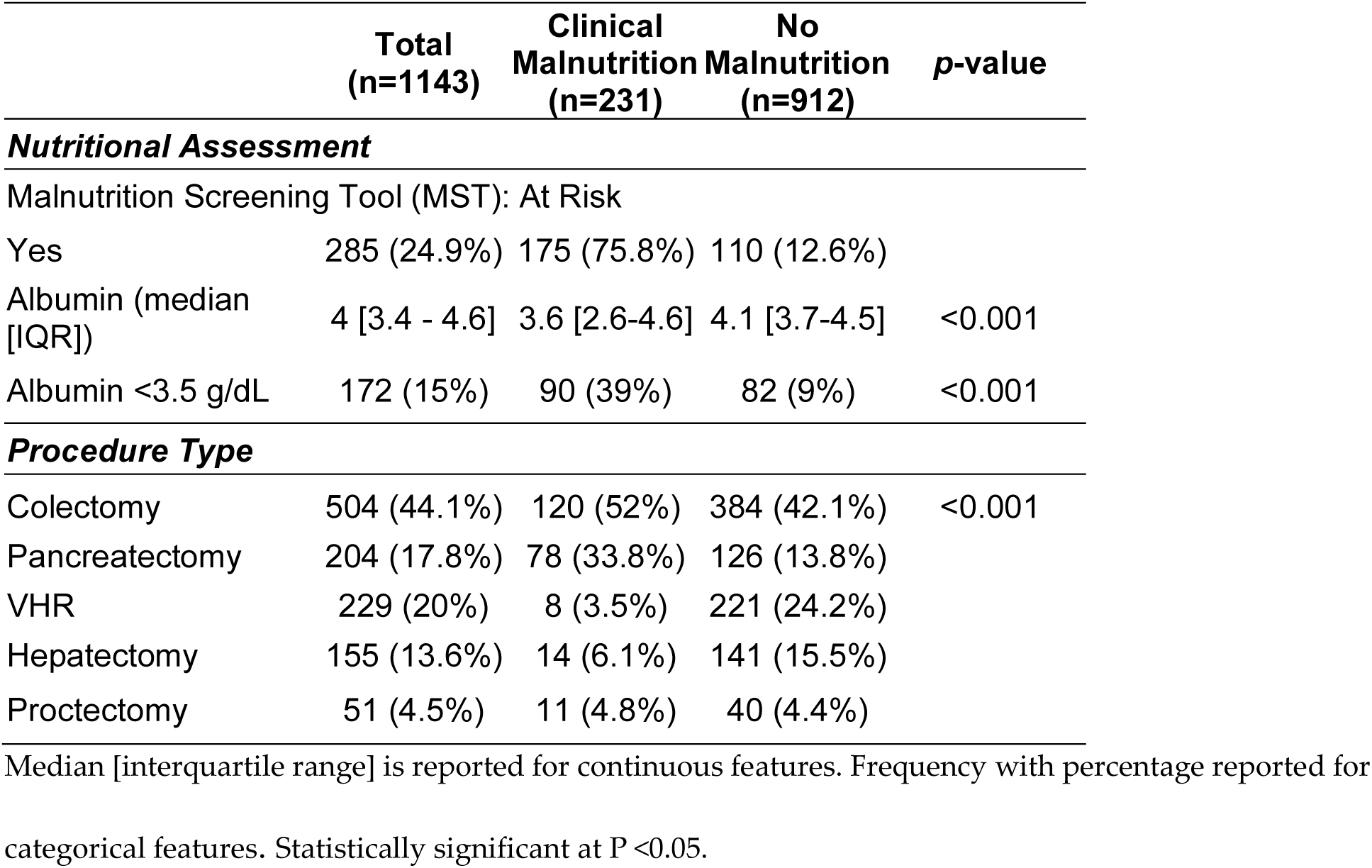
Nutritional Assessment and Procedure Type of Study Population.

#### Performance of Standard Screening

The Malnutrition Screening Tool (MST), administered within 24 hours of admission to the hospital following surgery, identified 285 patients (24.9%) as “at risk” for malnutrition (Table 2). Notably, 27.2% of patients ultimately diagnosed with clinical malnutrition did not screen positive on the MST but were identified postoperatively based on clinical indications for supplemental nutrition support.

### Predictive Model Development and Performance

Due to established sex-specific differences in body composition^25^, all models were developed separately for males and females. Three progressively comprehensive models were developed using an incremental approach to systematically evaluate the added value of imaging features (Table 3):

**Table 3.** Performance of models in predicting clinical malnutrition for Males (A) and Females (B).

| **Table 3A: Predictive performance of models in Males (N=544)** |  |  |  |  |  |
| --- | --- | --- | --- | --- | --- |
| Models | Features included (N) | Brier class | AUROC (95% CI) | p-value vs. Base Model | p-value vs. L3 Model |
| Base Clinical | 5 | 0.140 | 0.75 (0.70-0.80) | - | - |
| Base + L3 | 7 | **0.159** | 0.78 (0.74-0.83) | 0.027 | - |
| Base + L3 + Volumetric | 14 | **0.129** | 0.81 (0.77-0.85) | 0.004 | 0.024 |
| **Table 3B: Predictive performance of models in Females (N=599)** |  |  |  |  |  |
| Models | Features included (N) | Brier class | AUROC (95% CI) | p-value vs. Base Model | p-value vs. L3 Model |
| Base Clinical | 5 | 0.142 | 0.70 (0.65-0.75) | - | - |
| Base + L3 | 7 | 0.148 | 0.73 (0.68-0.78) | 0.059 | - |
| Base + L3 + Volumetric | 21 | 0.124 | 0.79 (0.75-0.84) | <0.001 | <0.001 |

Base clinical model contains (Age, weight, race, tobacco use and ASA classification). AUROC (area under the receiver operator characteristic curve) represents discriminative performance of model. Brier class reflects calibration performance of model. Delong test is used to compare the AUROC to determine if the difference in their discriminative performance is statistically significant. Statistically significant at $P < 0.05$ .

#### Model 1 – Base Clinical Model

Included preoperative clinical variables significantly associated with malnutrition on univariate analysis (age, race, weight, ASA class, and tobacco use). The base model achieved moderate discrimination with AUROC of 0.75 (95% CI: 0.70-0.80) for males and 0.70 (95% CI: 0.65-0.75) for females, with Brier scores of 0.140 and 0.142, respectively.

#### Model 2 – Base + L3 Imaging

Added traditional single-slice imaging features (L3 SMI and SMRA) to base clinical variables. This improved discrimination to AUROC of 0.78 (95% CI:0.74-0.83) for males and 0.73 (95% CI: 0.68-0.78) for females, with significant improvements compared to the base model for males (p=0.027) and a trend toward significance for females (p=0.06). However, Brier scores increased to 0.159 for males and 0.146 for females, indicating that while L3 features improved discrimination, they paradoxically worsened calibration.

#### Model 3 – Base + L3 + Volumetric Imaging

Incorporated comprehensive 3D body composition features (volume index and attenuation for five muscle groups and two fat depots) in addition to base clinical variables and L3 measurements. Elastic Net regularization selected 14 features for males and 21 features for females. This comprehensive model achieved the highest discrimination with AUROC of 0.81 (95% CI: 0.77-0.85) for males and 0.79 (95% CI: 0.75-0.84) for females; this comprehensive model showed significant improvement for males and females compared to both Model 1 (p=0.004 and p<0.00001, respectively) and Model 2 (p=0.02 and p=0.0001, respectively). Critically, the comprehensive model also achieved the best calibration with Brier scores of 0.129 for males and 0.124 for females—lower than both previous models—indicating that volumetric features improved both discrimination and the accuracy of probability estimates (Table 3). The incremental addition of imaging features progressively improved model performance for both males and females, as illustrated in AUROC curves (Figure 3).

**Figure 3:**
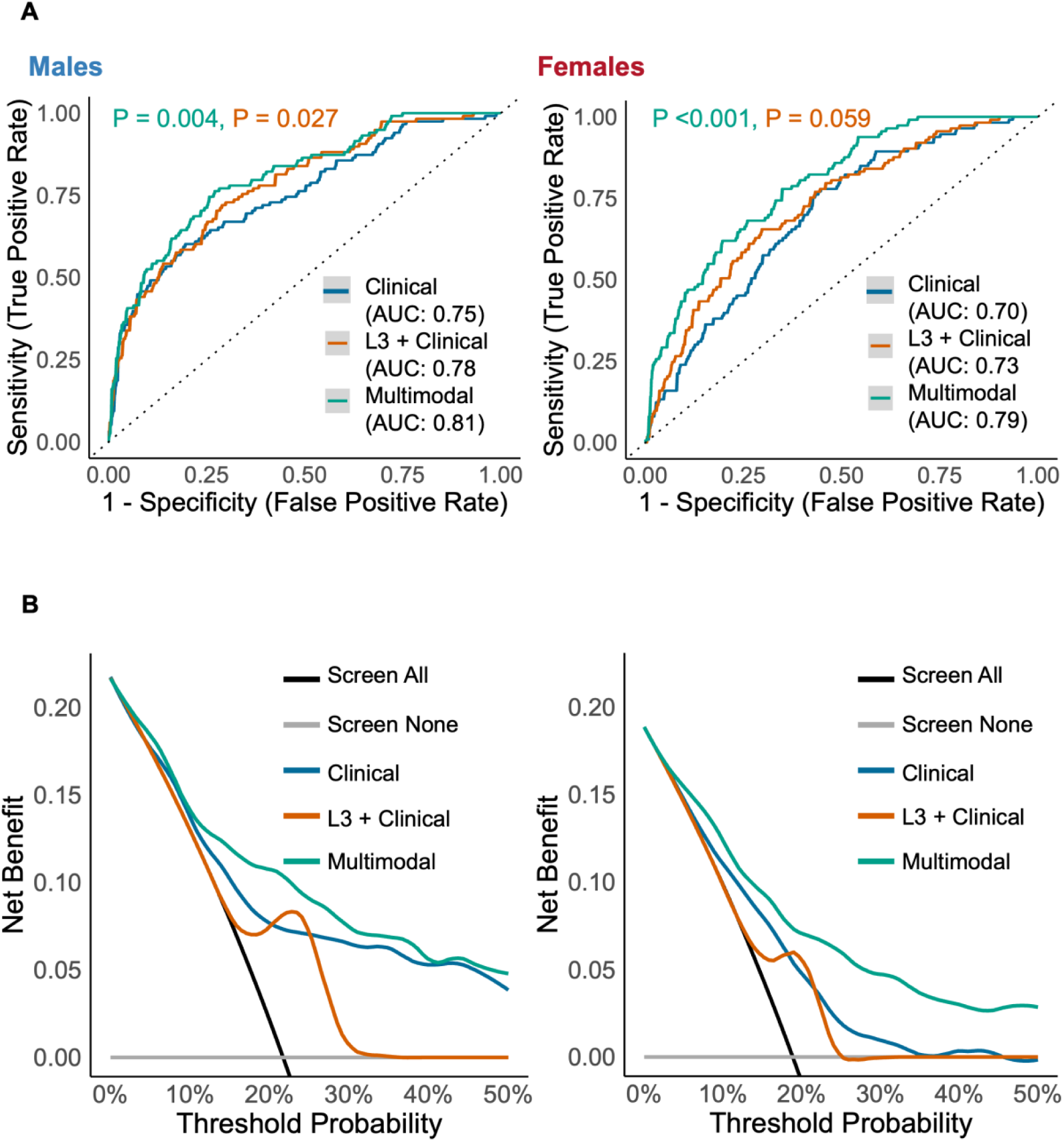
(A) ROC curves of Elastic Net regression models predicting clinical malnutrition for (i) Males and (ii) Females separately. Improvement in model performance is demonstrated with the addition of imaging features. ROC = receiver operating characteristic. (B) Decision curve analyses to determine net benefit for screening for clinical malnutrition in (i) Males and (ii) Females prior to abdominal surgery with proposed multimodal model

### Sex-Specific Feature Importance

Feature importance in the base clinical model demonstrated sex-specific patterns (Figure 4, top panels). For males, low body weight was the strongest predictor of malnutrition (β = -0.931, rank 1), followed by non-white race (β = -0.879, rank 2) and tobacco use (β = +0.411, rank 3). ASA classification (β = +0.282, rank 4) and age (β = +0.015, rank 5) had smaller contributions. For females, ASA classification dominated the model (β = +1.050, rank 1), with weight (β = -0.563, rank 2) and tobacco use (β = +0.226, rank 3) also contributing. Race (β = +0.170, rank 4) and age (β = +0.019, rank 5) had minimal impact, highlighting sex-specific risk profiles.

**Figure 4:**
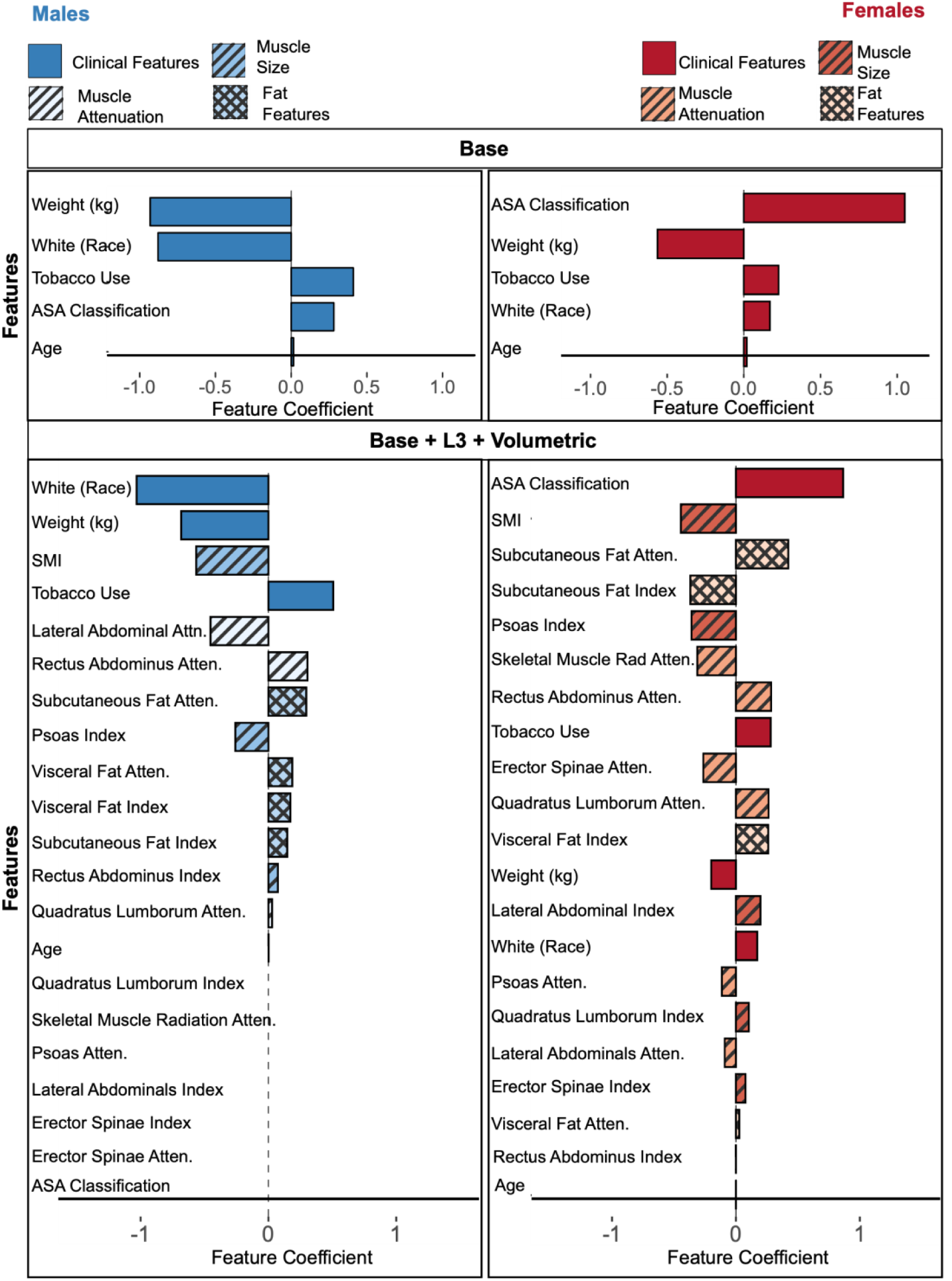
Feature importance for elastic net regression models determined by absolute of feature coefficients, stratified by sex. The top panel represents base model with the comprehensive base + L3 + volumetric model on the bottom. Positive coefficient indicates increase in log-odds of malnutrition. Negative coefficient represents inverse relationship and decreased log-odds of malnutrition.

Adding L3 imaging features shifted feature importance patterns (Model 2, supplemental figure 2). For males, race became the strongest predictor (β = -0.156, rank 1), followed by L3 SMI (β = - 0.099, rank 2), with weight (β = -0.097, rank 3) and tobacco use (β = +0.092, rank 4) having similar importance. L3 SMRA contributed modestly (β = +0.08, rank 6) and age (β =0.002, rank 7). For females, ASA classification remained the dominant predictor (β = +0.127, rank 1), with L3 SMI as the second most important feature (β = -0.083, rank 2). Weight (β = -0.057, rank 3), L3 SMRA (β = -0.032, rank 4), race (β = 0.032, rank 5), tobacco use (β = 0.020, rank 6) and age (β = 0.003, rank 7) also contributed.

The comprehensive imaging model (Model 3) incorporated 14 features for males and 21 features for females following Elastic Net regularization (Figure 4, bottom panels). Feature importance patterns differed dramatically between sexes, with striking differences in both the features retained and their relative importance (Table 4).

For males, race emerged as the overwhelmingly dominant predictor (β = -1.031, rank 1), substantially stronger than in the base clinical model (β = -0.879). Weight (β = -0.683, rank 2), L3 SMI (β = -0.565, rank 3), and tobacco use (β = +0.507, rank 4) were the next most important features. Among comprehensive body composition features, lateral abdominus attenuation was the strongest contributor (β = -0.454, rank 5), followed by rectus abdominus attenuation (β = +0.306, rank 6), subcutaneous fat attenuation (β = +0.296, rank 7), and psoas volume index (β = -0.259, rank 8). Fat depot features including visceral fat attenuation (β = +0.187, rank 9), visceral fat volume (β = +0.172, rank 10), and subcutaneous fat volume (β = +0.147, rank 11) contributed moderately. Notably, Elastic Net eliminated 7 features entirely, including ASA classification (β = 0), L3 SMRA (β = 0), and all erector spinae, lateral abdominus volume, psoas attenuation, and quadratus lumborum volume measurements, indicating these provided no incremental predictive value after accounting for the dominant demographic and selected body composition features.

For females, the comprehensive model retained all 21 features, indicating that the full spectrum of clinical, demographic, and body composition measurements contributed to prediction. ASA classification remained the dominant predictor (β = +0.865, rank 1), even stronger than in the base model. L3 SMI was the second most important feature (β = -0.445, rank 2), followed by subcutaneous fat attenuation (β = +0.422, rank 3). Unlike males, females showed important contributions from both fat volume measures—subcutaneous fat volume (β = -0.368, rank 4) and visceral fat volume (β = +0.261, rank 11)—alongside psoas volume (β = -0.358, rank 5). L3 SMRA, which was eliminated in males, contributed meaningfully for females (β = -0.312, rank 6). Multiple muscle attenuation features were retained: rectus abdominus (β = +0.284, rank 7), erector spinae (β = -0.263, rank 9), quadratus lumborum (β = +0.263, rank 10), psoas (β = -0.114, rank 15), and lateral abdominus (β = -0.090, rank 17). Clinical variables tobacco use (β = +0.281, rank 8) and weight (β = -0.200, rank 12) contributed moderately, while race (β = +0.172, rank 14) and age (β = -0.0001, rank 21) had minimal impact.

These sex-specific patterns reveal fundamental differences in malnutrition prediction. For males, race dominates independent of all body composition measurements, with Elastic Net eliminating ASA classification entirely despite its importance in the base model. The model selects only 14 of 21 possible features, favoring parsimony. For females, all 21 features contribute, with ASA classification remaining the strongest predictor across all three models. The retention of L3 SMRA for females but not males, and the broader array of muscle attenuation features in the female model, support the hypothesis that muscle quality assessment is more important for females than males. The distinct feature sets underscore the necessity of sex-stratified modeling for accurate malnutrition prediction.

### Clinical Utility and Decision Support Tools

Decision curve analysis demonstrated that the comprehensive imaging model provided superior net benefit compared to the base clinical model, L3-only model, and “treat all” or “treat none” strategies across a clinically relevant range of threshold probabilities (10-40%), supporting its utility for preoperative malnutrition screening (Figure 3b).

## Discussion

### Principal Findings

This study demonstrates that comprehensive, automated body composition analysis from routine preoperative CT scans can effectively predict clinical malnutrition in surgical patients, with progressive improvements achieved through incremental addition of imaging features. Four key findings emerged:

First, incorporating L3 single-slice imaging measurements significantly improved discrimination over clinical variables alone for both sexes (males: AUROC 0.78 vs 0.75, p=0.031; females: AUROC 0.73 vs 0.70, p=0.059), validating the utility of automated single-slice body composition assessment. However, L3 features paradoxically worsened model calibration despite improving discrimination (males: Brier score 0.1593 vs 0.1395; females: 0.1475 vs 0.1424), indicating that predicted probabilities became less accurate even as risk ranking improved.

Second, adding comprehensive volumetric features to the L3-enhanced model further improved discrimination significantly (males: AUROC 0.81, p=0.0019 vs L3 model; females: AUROC 0.794, p<0.00001 vs L3 model) and, critically, substantially improved calibration beyond even the base clinical model (males: Brier score 0.1286; females: 0.1244). This suggests that volumetric measurements provide corrective information that enhances both discrimination and the reliability of probability estimates.

Third, feature importance patterns differed substantially between sexes and evolved across models in striking ways. For males, race emerged as the overwhelmingly dominant predictor in the comprehensive model (β = -1.031, rank 1), substantially outweighing all body composition measurements and intensifying from its importance in the base model (β = -0.879, rank 2). Notably, ASA classification—which contributed to the base model—was eliminated entirely by Elastic Net in the comprehensive model (β = 0), as was L3 SMRA. The male model selected only 14 of 21 possible features. For females, ASA classification consistently dominated across all three models, achieving its strongest effect in the comprehensive model (β = +0.865, rank 1). Unlike males, the female model retained all 21 features, including L3 SMRA (β = -0.312, rank 6) and five distinct muscle attenuation measurements, supporting the hypothesis that muscle quality assessment across multiple sites is more important for females than males.

Fourth, decision curve analysis confirmed that the comprehensive imaging model provided superior net benefit across clinically relevant threshold probabilities, supporting its potential utility for preoperative screening.

Our findings build upon and extend prior research on body composition and surgical outcomes. Single-slice L3 measurements have been extensively studied as markers of sarcopenia and myosteatosis, with multiple studies demonstrating associations between these features and adverse outcomes after abdominal surgery. ^10–17,36,37^ However, translation to clinical practice has been limited by the need for manual segmentation, lack of automation, and inadequate incorporation into preoperative workflows. ^5,6,42^ Recent advances in deep learning have enabled both automated segmentation ^19–22,43–46^ and more comprehensive assessment that include volumetric measurements. Our approach extends this work by demonstrating that comprehensive volumetric assessment provides substantial additional benefit beyond L3 measurements. The incremental improvement from L3 to comprehensive models (males: ΔAUROC +0.03, p=0.0019; females: ΔAUROC +0.067, p=0.0001) indicates that evaluating multiple muscle groups and fat depots captures information not available from a single anatomical cross-section. This is particularly evident for females, where the comprehensive model showed more substantial improvements, consistent with recent findings by Xie et al. ^47^ who observed that muscle quality assessment across multiple sites better characterized malnutrition in older female adults.

Importantly, our calibration analysis reveals a previously unreported finding: L3 features, while improving discrimination, introduce calibration issues when added to clinical variables. This discrimination-calibration tradeoff has not been systematically examined in prior body composition studies, which have typically focused exclusively on AUROC or association measures. The finding that comprehensive volumetric features improve both discrimination and calibration—effectively “correcting” the calibration problems introduced by L3 alone—has important implications for clinical implementation and suggests that single-slice measurements may not adequately represent the heterogeneity of body composition changes during malnutrition.

Our study also reveals a striking and previously unexamined finding: the differential role of demographic versus clinical predictors by sex. Race emerged as the single most important predictor for males (β = -1.031), even more dominant than in models without imaging features, while having minimal impact for females (β = +0.172, rank 14). Conversely, ASA classification dominated female models (β = +0.865) but was eliminated entirely for males. This pattern suggests that malnutrition risk is not simply mediated through parallel pathways in males and females, but that sex-based metabolic differences may potentiate or amplify the nutritional sequelae of social determinants of health – a distinction with meaningful implications for how perioperative risk is assessed and addressed across diverse patient populations.

The high prevalence of malnutrition in our cohort (20.2% overall, 38.2% in pancreatic surgery) aligns with published rates ^4,48^ and underscores the clinical significance of this problem. Our preoperative prediction models—particularly the comprehensive model combining clinical variables, L3, and volumetric features—address this gap in both timing and sensitivity for surgical patients, creating opportunities for nutritional optimization before surgery when intervention is most effective.

### Clinical Implications and Translational Impact

Current practice relies on postoperative screening, which occurs only after patients have undergone surgical stress—too late for preemptive intervention. Preoperative malnutrition is a modifiable risk factor, ^1,30^ and accumulating evidence suggests that nutritional prehabilitation can improve surgical outcomes, particularly in high-risk populations. ^49,50^ However, without objective, scalable screening tools for the outpatient setting, identifying candidates for intervention remains challenging.

Our automated approach leverages CT scans already obtained for surgical planning, requiring no additional imaging, cost, or patient burden. The comprehensive segmentation algorithm runs automatically, extracting L3 and volumetric body composition features without manual input. ^23,24^ The comprehensive model, achieving both superior discrimination and optimal calibration, represents the preferred approach for clinical implementation. The well-calibrated probabilities enable clinicians to make informed decisions about intervention intensity based on accurate risk estimates. Decision curve analysis confirmed that implementing these screening models would provide net benefit compared to current practice (treat none) or universal intervention (treat all) across clinically relevant threshold probabilities.

### Sex-Specific Patterns and Biological Underpinnings

The marked sex differences in feature importance patterns observed across all three models reflect fundamental biological differences in body composition and nutritional physiology. Males and females differ in baseline muscle mass, fat distribution, hormonal regulation of metabolism and response to catabolic stress. ^51–54^ Our previous work demonstrated that males and females with malnutrition exhibit distinct body composition phenotypes, and the current prediction models confirm that these differences translate to divergent risk profiles requiring sex-stratified modeling.^25^

For males, race emerged as the overwhelmingly dominant predictor in the comprehensive model (β = -1.031, rank 1), substantially outweighing all other features including body composition measurements. ASA classification—contributing in the base model (β = +0.282)—was eliminated entirely in the comprehensive model. The parsimonious 14-feature model suggests malnutrition prediction in males relies primarily on demographic factors, with select body composition features (L3 SMI, lateral abdominus attenuation, subcutaneous fat attenuation, psoas volume) adding incremental value.

The dominance of race, which strengthens rather than attenuates with comprehensive body composition data, may reflect unmeasured socioeconomic factors (food security, healthcare access), referral patterns at this quaternary care center, healthcare utilization differences, or statistical artifacts including multicollinearity and suppression effects. Alternative explanations including disease severity at presentation, procedure type confounding, or sample-specific patterns cannot be excluded. External validation is critical to determine whether this represents generalizable disparity requiring intervention or site-specific artifact.

For females, the models demonstrated a markedly different hierarchy. ASA classification dominated all three models: base model (β = +1.050, rank 1), L3 model (β = +0.127, rank 1), and comprehensive model (β = +0.865, rank 1). This consistency demonstrates that comorbidity burden is the primary driver of malnutrition risk in females, independent of body composition. This contrasts sharply with males, where ASA was eliminated entirely in the comprehensive model. Further, race had minimal predictive value across all female models (base: β = +0.170, rank 4; L3: β = +0.032, rank 5; comprehensive: β = +0.172, rank 14). This striking sex difference—race dominant for males, negligible for females—suggests fundamentally different risk profiles that cannot be explained by biological factors alone.

The female comprehensive model retained all 21 features, indicating more complex, multifactorial pathophysiology. L3 SMI was consistently second most important, strengthening from β = -0.083 (L3 model) to β = -0.445 (comprehensive model). Notably, L3 SMRA—eliminated in males—contributed meaningfully for females (β = -0.312, rank 6). Subcutaneous fat attenuation emerged as the third strongest predictor (β = +0.422), highlighting adipose tissue remodeling as a key nutritional stress marker. Both fat volume and quality contributed: subcutaneous fat volume (β = -0.368, rank 4), visceral fat volume (β = +0.261, rank 11), and visceral fat attenuation (β = +0.027, rank 19).

The female model retained five muscle attenuation features (rectus abdominus, erector spinae, quadratus lumborum, psoas, lateral abdominus) compared to only three in males, supporting the hypothesis that muscle quality deterioration may be a more sensitive biomarker of malnutrition in women than muscle size alone. Females inherently have smaller muscle mass and higher intramuscular fat at baseline due to estrogen’s effects on adipose tissue distribution and metabolism. ^52–54^ During malnutrition, fatty infiltration of muscle (myosteatosis) may occur more readily or be more functionally significant in females. The impact of myosteatosis on risk is corroborated by a recently published paper by Kuchnia et al. that demonstrated increased mortality among both males and females with decreased muscle quality; importantly, they also found that muscle size was not predictive for females, highlighting the importance of including multiple body composition measures. ^55^

The positive coefficients for fat attenuation merit particular attention as they provide objective biomarkers of nutritional stress distinct from simple volume loss. Increased adipose tissue attenuation on CT has been linked to lipolysis, decreased lipid droplet size, increased fibrosis, edema, and adipose tissue remodeling seen in cachexia and former obesity. ^56–61^ Studies in cancer cachexia demonstrate that adipose tissue undergoes dramatic remodeling with increased attenuation reflecting cellular changes that precede frank weight loss, ^58^ and adipose tissue density (attenuation) independently predicts mortality in older adults. ^56^

The substantially different feature sets selected by Elastic Net—with only 14 features retained for males versus 21 for females, and virtually no overlap in the specific features and their relative importance—underscore the biological rationale for sex-stratified modeling. These models are not simply applying different weights to the same variables; they are fundamentally different predictive algorithms reflecting distinct pathophysiology of malnutrition in males and females.

### Limitations

Several limitations should be considered when interpreting these findings. First, as a single-center retrospective study, generalizability to other institutions, patient populations, and surgical contexts requires validation. Our cohort consisted of patients undergoing five specific abdominal operations at a quaternary care center; performance in other surgical specialties or community hospital settings remains unknown. The dominance of race in males—strengthening from base to comprehensive models—may reflect unmeasured confounding (socioeconomic status, food security, healthcare access), single-center referral patterns, disease severity differences at presentation, or statistical artifacts including multicollinearity and suppression effects. External validation examining racial distribution, procedure types, disease stage, and socioeconomic factors is essential to distinguish true associations from site-specific or statistical artifacts before drawing causal conclusions.

Second, the requirement for preoperative CT imaging within 90 days introduced selection bias toward patients with more complex disease or higher preoperative risk. While 65% of our cohort had ASA class ≥3, this may not represent all surgical candidates. Patients without preoperative CT scans were excluded, potentially missing lower-risk patients who might still benefit from screening.

Third, clinical malnutrition diagnosis by registered dietitians, while following standardized AAIM criteria, occurred postoperatively rather than preoperatively. ^33^ This timing discrepancy means we are predicting preoperative malnutrition risk but comparing it to postoperative clinical assessment of malnutrition. However, body composition changes slowly, and preoperative CT features likely reflect nutritional status at the time of imaging. Future studies incorporating preoperative dietitian assessment would strengthen the clinical validity of prediction models.

Fourth, our sample size (n=1,143 total; n=231 with malnutrition) limited statistical power for subgroup analyses by procedure type or age. The class imbalance (20% malnutrition prevalence) is representative of clinical reality but may affect model calibration.

Fifth, technical variability in CT scanning protocols (different vendors, tube voltages, contrast phases) may have introduced measurement noise, though our deep learning algorithm was designed for robustness across scanning conditions. The lack of external validation limits our ability to assess true model generalizability and calibration in new populations.

Sixth, the complex multivariate relationships revealed by Elastic Net regularization—particularly the paradoxical positive coefficients for some features (e.g., visceral fat volume index in both sexes, subcutaneous fat volume index in males)—highlight the challenge of interpreting individual feature contributions in high-dimensional models with correlated predictors. While these features collectively improve prediction, their individual coefficients may not align with univariate associations due to suppressor effects and multicollinearity. This underscores the importance of validating not just overall model performance but also examining whether feature importance patterns replicate in independent cohorts.

Finally, we did not compare our approach to other emerging nutritional screening tools or biomarkers (e.g. phase angle from bioelectrical impedance, serum markers of inflammation) that may complement imaging-based assessment. Integration of multiple data modalities may further improve prediction.

### Future Directions

Prospective validation in diverse, multicenter cohorts is essential to confirm model performance. Opportunistic CT screening approaches ^62^ could facilitate widespread implementation once validated. Interventional studies integrated with prehabilitation programs and enhanced recovery after surgery (ERAS) ^3^ are needed to demonstrate that screening-triggered nutritional interventions improve outcomes. Given distinct feature importance patterns, interventions may need sex-specific tailoring—targeting muscle quality in females versus addressing social determinants in males.

Extension to other patient populations (trauma, critical care, chronic disease) and integration with electronic health records for real-time clinical decision support could broaden impact. The growing availability of automated segmentation tools ^45^ and recognition of muscle mass as a vital sign ^6,7^ support widespread implementation feasibility.

## Conclusions

This study demonstrates that comprehensive, automated body composition analysis from routine preoperative CT scans significantly improves prediction of clinical malnutrition compared to clinical variables alone or traditional single-slice imaging measurements. Sex-specific models revealed distinct feature importance patterns reflecting biological differences in body composition and nutritional physiology, with females showing greater reliance on muscle quality and fat depot characteristics while males demonstrated stronger contributions from muscle size and demographic factors. The superior performance of volumetric imaging over L3 single-slice measurements support adoption of comprehensive assessment for accurate preoperative risk stratification. By identifying at-risk patients before surgery, this approach creates opportunities for targeted nutritional intervention when it is most likely to improve outcomes. Implementation of automated CT-based malnutrition screening could transform perioperative care by enabling objective, scalable identification of high-risk patients in the outpatient setting, ultimately improving surgical outcomes through timely nutritional optimization.

## Data Availability

All data produced in the present study are available upon reasonable request to the authors

## Acknowledgements

We wish to acknowledge with gratitude the Penn Clinical Nutrition Support Service registered dietitians for their instrumental role in collecting the data that underpins this research. Their expertise and dedication were vital in gathering comprehensive and accurate information necessary for our study. Special thanks to James C. Gee for his valuable contribution in development of CT segmentation tool for labeling abdominal muscle and fat depots.

Conception and study design: V.M.G., C.C., W.R.W. and G.D.W. Data acquisition: V.M.G., R.A.D., W.R.W. and G.D.W., J.R. Data analysis and interpretation: R.A.D. S.V., R.S., C.C., Y.R., H.L., G.D.W., J.D. and V.M.G. Writing–drafting the article: V.M.G., R.A.D., S.V., W.R.W. and G.D.W. Writing—critical review and editing: V.M.G., S.D., G.V., R.K., C.C., H.S., G.D.W. and W.R.W. All authors have read and agreed to the published version of the manuscript.

## Data Availability

Data described in the manuscript, code book, and analytic code will be made available upon request pending application and approval

## Declaration of Generative AI and AI-assisted technologies in the writing process

During the preparation of this manuscript, the author(s) used Claude 3.5 Sonnet (Anthropic PBC, San Francisco, CA, USA) and Gemini 1.5 Pro (Google LLC, Mountain View, CA, USA) to improve the clarity, syntax, and grammatical flow of the text. After using this tool, the author(s) reviewed and edited the content as needed and take(s) full responsibility for the accuracy and integrity of the final publication.

## Abbreviations

The following abbreviations are used in this manuscript

BMI: Body Mass Index
CT: Computed Tomography
HU: Hounsfield Units
SMI: Skeletal Muscle Index
SMRA: Skeletal Muscle Radiation Attenuation
L3: 3rd Lumbar Vertebrae
AAIM: Academy ASPEN Indicators of Malnutrition
NSQIP: National Surgical Quality Improvement Program
OR: Odds Ratio
AUROC: Area under receiver operator characteristic curve
DCA: Decision curve analysis

## Supplementary Materials

**Supplemental Table 1.** List of imaging-derived body composition features.

|  |  |  |
| --- | --- | --- |
|  | Size Feature | Attenuation Feature (HU) |
| 2D | L3 Skeletal Muscle Index (cm<sup>2</sup>/m<sup>2</sup>) | L3 Skeletal Muscle Radiation Attenuation |
| 3D | Psoas Index (cm<sup>3</sup>/m<sup>2</sup>) | Psoas Attenuation |
| 3D | Quadratus Lumborum Index (cm<sup>3</sup>/m<sup>2</sup>) | Quadratus Lumborum Attenuation |
| 3D | Erector Spinae Index (cm<sup>3</sup>/m<sup>2</sup>) | Erector Spinae Attenuation |
| 3D | Rectus Abdominus Index (cm<sup>3</sup>/m<sup>2</sup>) | Rectus Abdominus Attenuation |
| 3D | Lateral Abdominals Index (cm<sup>3</sup>/m<sup>2</sup>) | Lateral Abdominals Attenuation |
| 3D | Visceral Fat Index (cm<sup>3</sup>/m<sup>2</sup>) | Visceral Fat Attenuation |
| 3D | Subcutaneous Fat Index (cm<sup>3</sup>/m<sup>2</sup>) | Subcutaneous Fat Attenuation |

**Supplementary Figure 1:**
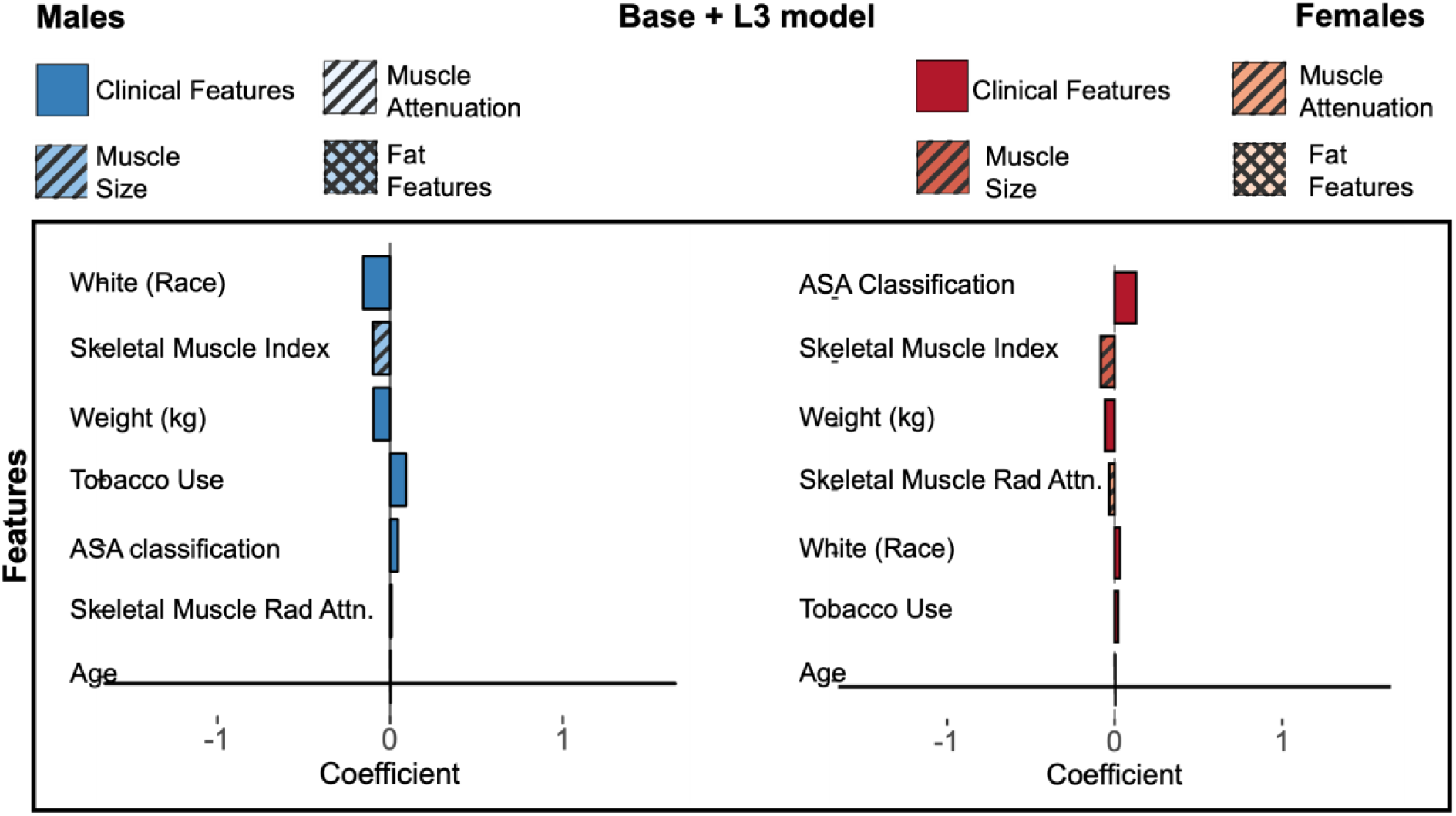
Feature importance plot for Base Clinical+L3 model.

**Supplementary Table 2:** Feature Coefficients in Final Logistic Regression Models including Volumetric imaging and Clinical features by Sex

| Features | $\beta$ Coefficient<br>(Males) | Importance Rank<br>(Males) | $\beta$ Coefficient<br>(Females) | Importance Rank<br>(Females) |
| --- | --- | --- | --- | --- |
| ***Clinical*** |  |  |  |  |
| Race | -1.031 | 1 | 0.172 | 14 |
| Weight | -0.683 | 2 | -0.200 | 12 |
| Tobacco Use | 0.507 | 4 | 0.281 | 8 |
| ASA class | - | - | 0.865 | 1 |
| Age at Time of Surgery | 0.002 | 14 | -0.0001 | 21 |
| ***L3 Features*** |  |  |  |  |
| Skeletal Muscle Index | -0.565 | 3 | -0.445 | 2 |
| Skeletal Muscle Radiation Attenuation | - | - | -0.312 | 6 |
| ***Muscle Volume Index*** |  |  |  |  |
| Psoas | -0.259 | 8 | -0.358 | 5 |
| Lateral Abdominals | - | - | 0.198 | 13 |
| Rectus Abdominus | 0.074 | 12 | -0.001 | 20 |
| Erector Spinae | - | - | 0.077 | 18 |
| Quadratus Lumborum | - | - | 0.104 | 16 |
| ***Muscle Attenuation*** |  |  |  |  |
| Psoas | - | - | -0.114 | 15 |
| Lateral Abdominals | -0.454 | 5 | -0.090 | 17 |
| Rectus Abdominus | 0.306 | 6 | 0.284 | 7 |
| Erector Spinae | - | - | -0.263 | 9 |
| Quadratus Lumborum | 0.029 | 13 | 0.263 | 10 |
| ***Fat Volume Index*** |  |  |  |  |
| Subcutaneous | 0.147 | 11 | -0.368 | 4 |
| Visceral | 0.172 | 10 | 0.261 | 11 |
| ***Fat Attenuation*** |  |  |  |  |
| Subcutaneous | 0.296 | 7 | 0.422 | 3 |
| Visceral | 0.187 | 9 | 0.027 | 19 |

