## Supplementary figures and images for "Development of an automated, imaging-based preoperative screening model for early identification of malnutrition in an abdominal surgery cohort"

### Supplemental Figure 1

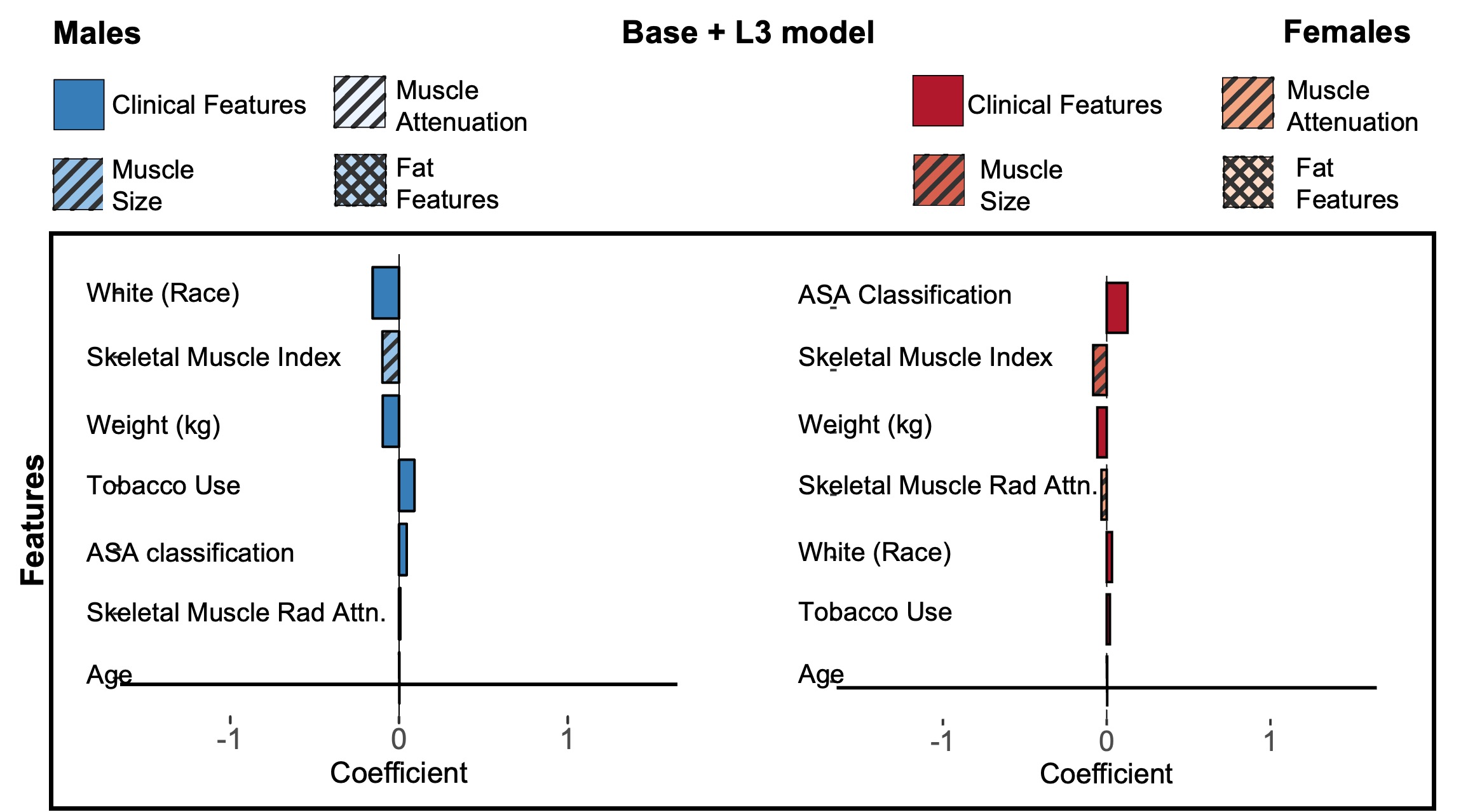
